# Reducing PIA by over 50% While Generating New Patient Flows: A Comprehensive Assessment of Emergency Department Redesign

**DOI:** 10.1101/2024.05.22.24306395

**Authors:** Opher Baron, Marko Duic, Dmitry Krass, Tianshu Lu, Zhoupeng Jack Zhang

## Abstract

**Background:** On June 6, 2011 the Emergency Department (ED) at Southlake Regional Health Center, a very high-volume ED, initiated a comprehensive redesign project to improve patient waiting times. The primary initial goal of the project was to reduce Time to Physician’s Initial Assessment (TPIA) - one of the Key Performance Indicators (KPIs) tracked by the Ontario Ministry of Health and Long-Term Care. The objective was to achieve a significant improvement in TPIA without sacrificing performance on any other important KPIs such as Length of Stay (LOS), Left Without Being Seen (LWBS), or time to admission (T2A). The effect on TPIA was immediate and dramatic: the 90−th percentile TPIA declining from 4 hrs to under 2.5 hrs, with further improvements seen over time. The patient in-flows also increased; anecdotally this increase was directly related to shorter wait time. However, like any other large-scale and on-going system redesign project, the impacts are not limited to the listed KPIs, but are multi-dimensional, affecting patient inflows, flows within the ED, workloads, staffing levels, etc. Thus, teasing out the impact of system redesign requires from other concurrent factors (population changes, staffing changes, etc.) requires a comprehensive system assessment. The available data exhibits auto-correlations, heteroscedasticity, and interdependence among variables, rendering simple statistical analysis of individual KPIs inapplicable. We develop a novel methodology and conduct counterfactual analysis demonstrating that the decrease in TPIA, as well as new patient inflows can indeed be attributed to the ED redesign. This suggests that a similar system redesign should be considered by other EDs looking to improve wait times.

**Objectives:** To (1) statistically estimate the impacts of the redesign project on various performance measures over time, (2) examine whether the project’s initial goal of improvement in TPIA without compromising other service performance measures was achieved, and (3) study whether the project impacted patient inflows.

**Methods:** We (1) estimate simultaneous equations models to quantify interdependent and timevarying relations among variables, (2) conduct an iterative counterfactual analysis to estimate the mean-level impacts of the project, and (3) construct 95% confidence intervals for the estimated impacts using the Bootstrap method.

**Results:** We study project impacts over 720 days after it was initiated. During this time, the 90*^th^* percentile of TPIA has been reduced by nearly 2.5 hours on average (translating into an over 50% improvement), with continuous improvement over the study period. This effect is statistically and operationally significant. The project also improved LOS for non-admitted patients (both acute and non-acute), and did not have statistically significant impact on LOS for admitted patients. There was also a decrease in LWBS, though it was not statistically significant. Thus the project achieved its stated primary goals. We also observed an increase in inflows of both acute and nonacute patients; our analysis confirms that this increase can be attributed to the project, indicating that improvements in TPIA attracted new patients to the ED. All of these effects have persisted over the 720-day post-project period.

**Conclusions:** The redesign project has significantly reduced TPIA over time while also improving some LOS measures; none of the waiting time KPIs were compromised. The reduction in TPIA also attracted significant volumes of new patients. However, the redesigned process was able to deal with this volume without compromising performance. The redesign project involved a number of major changes in ED operations. We provide an overview of these changes, and while our analysis cannot attribute specific project impacts to specific changes, we believe that implementing similar changes should receive strong consideration by other EDs.

**Conflicts of interest:** None

## Introduction

The long ED waiting time has been a significant problem in Canada over at least the past two decades. Evenson (2012) shows that among 11 countries, Canada has also the highest percentage of waiting longer than four hours before being treated. With an aging population and longer life expectancy, this trend of long delays in EDs is common in many countries and is also expected to continue if the situation would be left without proper intervention.

The ED at Southlake Regional Health Center in Newmarket, Ontario (Southlake RHC), is classified as a “very high volume ED” by the Ontario Ministry of Health and Long-Term Care (MOHLTC). In June 2011 the Southlake ED Management team launched a comprehensive redesign project (“project”) aimed at improving patient wait times. The primary goal of the project was to improve Time until Physician Initial Assessment (TPIA) without any adverse effect on other key Performance Indicators (KPIs) such as length of Stay (LOS) measures. Some background details of the ED redesign, as well as the choice of the primary goals, are provided below.

The primary purpose of the current paper is to conduct a comprehensive assessment of the impact of the project. We develop a novel methodology that can be adapted for evaluating the impacts of other large-scale institutional projects. There are several complicating factors in performing such an evaluation:

- While the project may have a definite start date (in the case of Southlake ED it was 6/6/2011), it often does not have an end date - rather, continuous improvements and adjustments are made as the project progresses. Thus, the evaluation must incorporate the evolution of the project over time.
- The project has a broad impact on the system and thus on all of its KPIs. Thus, the evaluation must incorporate the evolution of the project on several KPIs simultaneously.
- A large system-wide project affects multiple systems and performance measures, both internal and external (e.g., patient inflows). These effects create complex feedback loops with both positive and negative consequences. For example, reduction in wait times may attract more patients to the ED (anecdotally, this was the case at Southlake ED), which creates overcrowding and may increase wait times (i.e., TPIA and LOS) - a negative feedback. However, this could also trigger new improved efficiency initiatives implemented as part of the long-term redesign project to respond to higher patient volumes, which may lower the wait time - a positive feedback. Moreover, the profile of patients (e.g., ratio of Acute vs Non-Acute patients) visiting the ED may change over time, both due to feedback described above, and due to external reasons (e.g., demographic changes in the hospital’s catchment area). This may impact per-patient workloads.

Thus, simple statistical techniques (e.g., *t*−test) focusing on “before” and “after” comparisons of individual performance measures are insufficient. While simple analysis based on data descriptions in Table 2 suggests that the project was indeed successful (TPIA fell from 4.1 hours to 1.5 hours, while all LOS measures saw modest declines), we need to establish whether these effects were statistically significant while taking into account the dynamic changes, both internal and external, that occurred during the evaluation period. Thus, we need to model the entire dynamically evolving system and then to construct the “counterfactual trajectory” for this system: how the system would have evolved in the absence of the project. Moreover, we need to establish that the difference between the actually observed and the counterfactual trajectories is statistically significant with respect to the key measures of interest.

In this paper we outline such a methodology and apply it to the project at Southlake ED. Our methodology is based on simultaneous equation modeling (SEM) with time-series components. We estimate the model using the multi-stage least square approach (LeMay 1990) to capture the feedback effects among variables of interest. Using the estimated model, we conduct an iterative counterfactual analysis to quantify the short-run and long-term impacts of the project. Finally, we develop a multi-stage Bootstrap procedure to assess the statistical significance of the estimated impacts. We use these results to show that the Southlake ED project was, indeed, a success and the tangible improvements in the TPIA are due to the project and not some external factors. We also uncover a comprehensive set of impacts that the project had - from patient inflows to various LOS measures. In particular, our results indicate the improvements in TPIA did serve to attract significant new patient volumes to the hospital. However, the redesigned system was able to handle these additional volumes without adverse impacts on any of the KPIs.

The methodology we develop is quite general and can be applied to other large-scale redesign projects. Such comprehensive evaluation is particularly important when the project in question makes radical changes to the underlying system and may thus cause certain political resistance within the organization; the ability to attribute improvements in performance to the project and rule out external factors is especially valuable in such cases.

### ED Process and Project Background

The overall process at Southlake ED, which is typical of most EDs, is illustrated in Figure 1 (this is a stylized representation that only includes typical key steps). The “Acute” and “Non-Acute” patients are classified according to *Canadian Triage and Acuity Scale* (CTAS), see Bullard et al. (2017) for the current version. *Acute* corresponds to CTAS scores of 1, 2 or 3, and *Non-Acute* to CTAS scores of 4 or 5. The key performance measures are indicated in the figure. These include time from triage until initial physician assessment (TPIA), patients who went through triage, but then left without being seen (LWBS), i.e., departed prior to the initial assessment, triage-to-admit decisions time (T2A, applies for admitted patients only), and length of stay (LOS) measured separately for Admitted (LOS*^Ad^*), Acute Non-Admitted (LOS*^Act NAd^*), and Non-Acute patients (LOS*^NAct^ ^NAd^*). While admissions for Non-Acute patients do happen, they are rare and not tracked separately.

**Figure 1.**
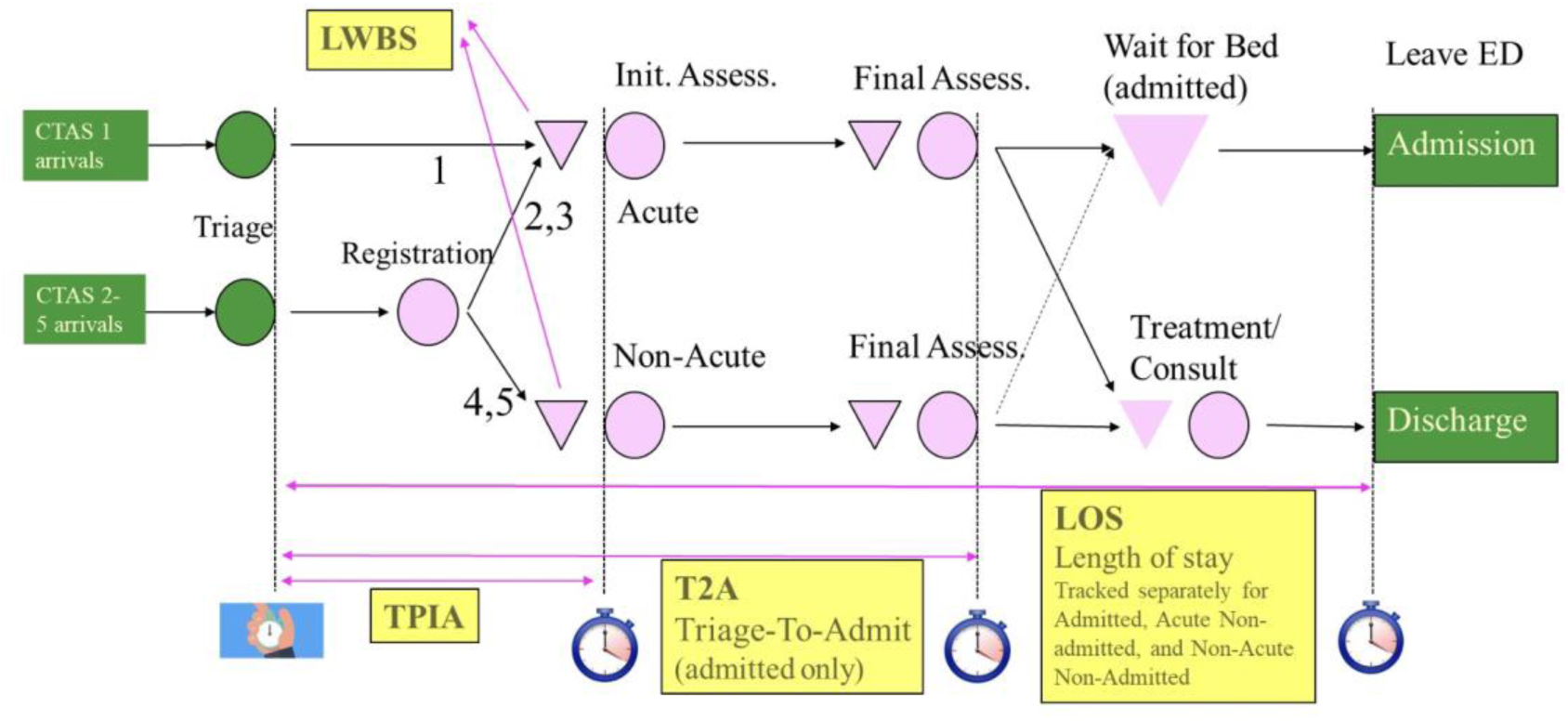
Diagram of the ED Process at Southlake. Waiting areas are represented by triangles. Prior to the projects, the main waiting area was in front of Triage.

The ED re-design project described in the current paper was, in part, motivated by the Pay-for-Results program (P4R) initiated by Ontario Ministry of Health and Long-Term Care (MOHLTC) in 2008, which provides financial incentives for hospitals to improve their performance (see, MOHLTC 2010, as well as Li et al. 2014, Vermeulen et al. 2016). Some of the relevant KPIs used by P4R include: TPIA, LOS*^Ad^*, LOS*^Act NAd^*, and LOS*^NAct NAd^*. All KPIs are measured at the daily 90*^th^* percentile level (not that some important KPIs, such as LWBS and T2A are not part of the P4R program). The P4R program rewards EDs for both, achieving top rank with respect to a given KPI relative to other provincial EDs of the same size, and for year-over-year improvement on each KPI.

Prior to the initiation of the project in 2011, Southlake ED had experienced long patient wait times - placing it in line with provincial averages for ED of this size, but considered unacceptable by hospital management. For example, the 90*^th^* percentile of TPIA was 4.1 hours (see Table 2).

On June 6*^th^*, 2011 the Southlake ED Management team launched a comprehensive redesign project aimed at improving patient wait times. Southlake’s ED redesign project’s primary goal was *to improve TPIA without adverse effects on any of the LOS measures*; it was expected that LWBS, which was hypothesized to be primarily driven by long TPIA, would also improve should the project be successful. The primary focus on TPIA was due to several factors. First, the period between triage and physician initial assessment is viewed by many emergency medicine practitioners as the most dangerous part of a patient’s visit to the ED, both due to potential triage errors, and due to lack of monitoring during this period: should the patient’s condition progress while waiting for the initial assessment, the health outcome may be poor. Indeed, long TPIA is associated with medical complications or even fatalities (see Linnane 2020, Smith and Quon 2019). As described in Ang et al. (2016), TPIA affects both ambulance routing for acute patients and ED choice for sub-acute patients. Second, it was felt that TPIA was entirely within the control of ED, while other KPIs were affected by external factors (e.g., LOS may be impacted by long waits for consultation with specialists). Third, should the initial stage of the project prove successful, it was felt that LOS measures could be addressed in later stages.

The project involved a number of fundamental operational changes in the ED department. These changes, summarized on Table 1, ranged from re-configuring the physical layout of the ED (the waiting area in front of the registration and triage desks where most patients were waiting prior to the project was eliminated) to the change in the design and scheduling of physician’s shifts, the introduction and heavy use of “navigators” (non-medical personnel, usually students, whose duties ranged from making sure all documentation and reports were ready prior to physician entering the examination room to routing the physician to the next patient to be seen). See Whatley (2016) for further description of these changes.

**Table 1.**
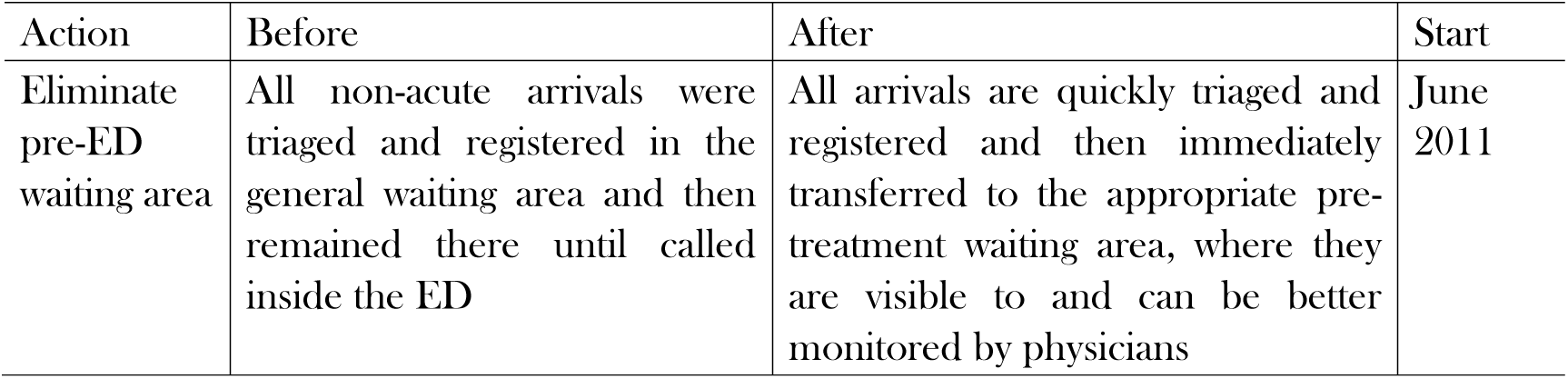
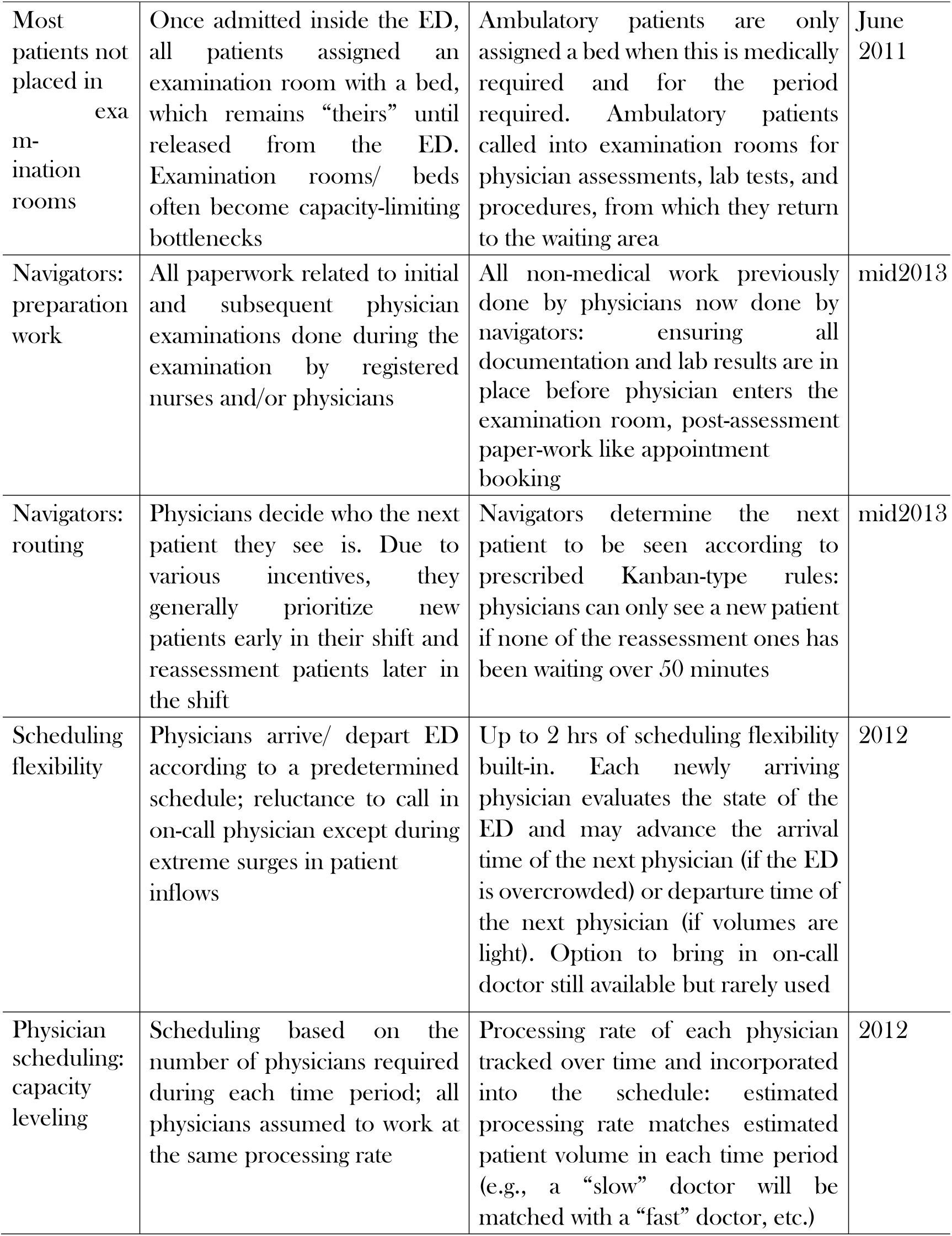
Brief Description of ED Redesign Project in Southlake RHC.

| Action | Before | After | Start |
| --- | --- | --- | --- |
| Eliminate pre-ED waiting area | All non-acute arrivals were triaged and registered in the general waiting area and then remained there until called inside the ED | All arrivals are quickly triaged and registered and then immediately transferred to the appropriate pre-treatment waiting area, where they are visible to and can be better monitored by physicians | June 2011 |
| Most patients not placed in examination rooms | Once admitted inside the ED, all patients assigned an examination room with a bed, which remains “theirs” until released from the ED. Examination rooms/ beds often become capacity-limiting bottlenecks | Ambulatory patients are only assigned a bed when this is medically required and for the period required. Ambulatory patients called into examination rooms for physician assessments, lab tests, and procedures, from which they return to the waiting area | June 2011 |
| Navigators: preparation work | All paperwork related to initial and subsequent physician examinations done during the examination by registered nurses and/or physicians | All non-medical work previously done by physicians now done by navigators: ensuring all documentation and lab results are in place before physician enters the examination room, post-assessment paper-work like appointment booking | mid2013 |
| Navigators: routing | Physicians decide who the next patient they see is. Due to various incentives, they generally prioritize new patients early in their shift and reassessment patients later in the shift | Navigators determine the next patient to be seen according to prescribed Kanban-type rules: physicians can only see a new patient if none of the reassessment ones has been waiting over 50 minutes | mid2013 |
| Scheduling flexibility | Physicians arrive/ depart ED according to a predetermined schedule; reluctance to call in on-call physician except during extreme surges in patient inflows | Up to 2 hrs of scheduling flexibility built-in. Each newly arriving physician evaluates the state of the ED and may advance the arrival time of the next physician (if the ED is overcrowded) or departure time of the next physician (if volumes are light). Option to bring in on-call doctor still available but rarely used | 2012 |
| Physician scheduling: capacity leveling | Scheduling based on the number of physicians required during each time period; all physicians assumed to work at the same processing rate | Processing rate of each physician tracked over time and incorporated into the schedule: estimated processing rate matches estimated patient volume in each time period (e.g., a “slow” doctor will be matched with a “fast” doctor, etc.) | 2012 |

As noted in Table 1, at the commencement of the project, two changes were implemented immediately. The first one was the new ED layout, where instead of sitting in the waiting area outside of ED patients were immediately admitted inside, and waited in the central area surrounded by examination rooms. The main effect was to make the “workload” of the waiting patients visible to the ED physicians; this leveraged a well-documented effect in behavioral queuing literature that human “servers” tend to speed up in response to workload; see e.g., Do et al. (2018).

The second change related to the treatment of ambulatory patients of all priority classes. Whereas prior to the project the standard practice was to allocate an ED bed where the patient would remain until discharge, the new practice was to only assign a bed when medically required. All other ambulatory patients were called into examination rooms for physician assessments and other medical procedures, but would spend the remaining time in waiting room chairs. The primary effect of this change was to improve patient flows which were frequently blocked by unavailability of beds prior to the project. It also made it easier to stock and operate the examination rooms efficiently. These two changes resulted in over 1-hr reduction of 90*^th^* percentile of TPIA within days of the project start.

The remaining changes were implemented gradually, as indicated in the last column of Table 1; many adjustments were (and continue to be) made on on-going basis. The overall success of the project was heavily based on the buy-in by all members of the ED leadership team, who met daily to identify impediments to smooth patient flows within the ED and ways to mitigate them. Many of the principles they followed are based on the Lean Operations/ Toyota Production System. (See Ng et al. (2010) for a description of another successful implementation of these principles in a Canadian medical setting).

By 2016, Southlake RHC ranked in first place in TPIA among 73 EDs in Ontario, with a median 90*^th^* percentile of TPIA of 1.4 hours, compared to the province average of 3.3 hours. As seen in Table 2, there were improvements in LWBS and small-scale improvements in LOS measures. These improvements were achieved while the inflows of Acute and Non-Acute patients increased by over 30%. As documented in the remainder of the current paper, these improvements can, with a high degree of certainty, be attributed to the project, rather than background factors or increases in medical resources.

**Table 2.** Summary Statistics. The TPIA, and LOS are measured in hours at the 90−th daily percentile level. T2A is measured in hours at the daily mean. Acute, Non-Acute, ADM and LWBS are daily average patient counts. DayPhysician and NightPhysician are in hours.

|  |  | Min | Median | Mean | Max | Std. error |
| --- | --- | --- | --- | --- | --- | --- |
| TPIA | Overall | 0.75 | 1.6 | 2.26 | 8.43 | 1.38 |
|  | Before | 1.5 | 4 | 4.1 | 8.4 | 1.2 |
|  | After | 0.75 | 1.42 | 1.51 | 3.64 | 0.39 |
| T2A | Overall | 5.2 | 10 | 10.4 | 22 | 2.4 |
|  | Before | 6 | 10.3 | 10.5 | 16.1 | 2.4 |
|  | After | 5.18 | 9.99 | 10.38 | 21.97 | 2.38 |
| LOS<sup>Ad</sup> | Overall | 8.6 | 27.2 | 27.5 | 73.5 | 8.4 |
|  | Before | 12.9 | 28.5 | 30.2 | 52.1 | 9.9 |
|  | After | 8.55 | 27.17 | 27.4 | 73.54 | 8.37 |
| LOS<sup>Act, NAd</sup> | Overall | 4.58 | 6.62 | 6.65 | 11.91 | 0.82 |
|  | Before | 5.6 | 7.3 | 7.6 | 11.9 | 1.2 |
|  | After | 4.58 | 6.58 | 6.61 | 10.92 | 0.78 |
| LOS<sup>NAct, NAd</sup> | Overall | 1.7 | 3.26 | 3.34 | 7.17 | 0.66 |
|  | Before | 2.2 | 3.4 | 3.6 | 7.2 | 0.8 |
|  | After | 1.7 | 3.26 | 3.33 | 6.58 | 0.65 |
| LWBS | Overall | 0 | 2 | 3.4 | 29 | 3.9 |
|  | Before | 0 | 6 | 7 | 29 | 5.2 |
|  | After | 0 | 2 | 1.97 | 11 | 1.78 |
| Acute | Overall | 95 | 163 | 164 | 278 | 28 |
|  | Before | 95 | 136 | 137 | 207 | 17 |
|  | After | 103 | 174 | 175 | 278 | 24.27 |
| NonAcute | Overall | 32 | 87 | 89 | 172 | 21 |
|  | Before | 36 | 70 | 71 | 130 | 13 |
|  | After | 32 | 95 | 95.66 | 172 | 19.27 |
| ADM | Overall | 8 | 29 | 29.2 | 55 | 6.3 |
|  | Before | 10 | 26 | 26.4 | 44 | 5.5 |
|  | After | 8 | 30 | 30.26 | 55 | 6.23 |
| DayPhysician | Overall | 0 | 27 | 27 | 36 | 3 |
|  | Before | 0 | 27 | 26.7 | 32.5 | 2.8 |
|  | After | 19 | 25 | 27.33 | 36 | 3.04 |
| NightPhysician | Overall | 0 | 13 | 13.4 | 18 | 1.8 |
|  | Before | 0 | 16 | 15.1 | 18 | 2.2 |
|  | After | 3 | 13 | 12.72 | 16 | 1.09 |

### Research Hypotheses

Our goal is to systematically evaluate the effectiveness of the Southlake ED redesign project. To this end we study the following research hypotheses:

**Hypothesis 1.** The redesign project has led to a significant reduction of the 90^th^-percentile of TPIA. In particular, this positive impact has persisted over time.

Hypothesis 1 is motivated by both on-site observations (as noted earlier, the TPIA dropped by over an hour almost immediately after the project start) and the before-and-after comparisons in Table 2.

**Hypothesis 2** The redesign project has directly led to an increase in patient arrivals to the ED.

This hypothesis is motivated by anecdotal evidence, where doctors in the catchment area of the ED were apparently actively advising their patients to visit the ED as a way to get faster access to imaging and specialists.

**Hypothesis 3.** The LOS measures did not increase for Admitted, Acute Non-Admitted, or NonAcute Non-Admitted groups as a result of the project.

This hypothesis reflects one of the primary goals of the project. There was a concern that improvement in TPIA would come at the expense of increasing LOS for some patient groups. Indeed, by having the physicians always prioritize new arrivals over patients already in the ED it is not hard to achieve a reduction in TPIA with the likely increase in LOS. The goal of the project was to achieve tangible improvements in TPIA via fundamental improvements to system operations rather than trading off TPIA vs LOS.

**Hypothesis 4** *Project led to a decrease in LWBS*.

It is widely assumed that LWBS is primarily driven by long TPIA, so this is a natural consequence of Hypothesis 1.

### Literature Review

Our work relates to the empirical literature on waiting time reduction in ED. Researchers have studied methods to reduce waiting time at different stages of patient flow, i.e., flow into, within, and out of ED (Saghafian et al. 2015, Anderson et al. 2023, Brouillette 2024). We particularly focus on waiting time within and out of ED. Some research in this literature compares waiting time before and after some intervention without computing standard deviations or constructing confidence intervals for average changes in waiting time (see, e.g., Jeong et al. 2014). Other research generally uses two approaches to compute confidence intervals. First is to use the standard deviations given by hypothesis testing methods such as Student’s t-test (Morais Oliveira et al. 2018). Second is based on logistic regression. The authors that adopt this approach typically discretize their measures for waiting time for logistic regression and then compute changes in the odds ratios with confidence intervals (Mataloni et al. 2018). Some work does not discretize time measures but uses generalized linear models to compute confidence intervals (see, e.g., Kawano et al. 2014). We find these approaches incomplete since by focusing on single-equation models they do not consider dynamic interactions and feedbacks between various parts of the system. Modeling such effects seems essential when evaluating a complex system redesign project.

Regarding the literature on simultaneous equation estimation (Hendry 1976, Hausman 1983), our work contributes to the estimation and statistical analysis of SEM with general error covariance structures; see Fair (1970) for the pioneering work on SEM estimation with auto-correlated data and Anderson et al. (2017) for recent progress on estimation methodology that accommodates both error auto-correlation and heteroscedasticity. In particular, LeMay (1990) develops a Multi-stage Least Square technique that not only applies to error terms with aforementioned features but also tackles the endogeneity issue in an equation system. Building upon LeMay (1990), the estimation procedure we propose extends to error auto-correlation of general orders. More importantly, we develop an iterative counterfactual estimator and a multi-stage Bootstrap procedure to construct non-parametric counterfactual intervals using techniques for time-series Boostrapping (see, e.g., Freedman and Peters 1984, Li and Maddala 1996, Davidson and Monticini 2014).

## Methods

### Data

We use a dataset from June 1*^st^*, 2009 to June 2*^nd^*, 2016, roughly corresponding to 2 years prior to the project start through the end of the initial stage of the project (after 2016 the objectives gradually shifted to reduction in LOS, while maintaining reduction in TPIA). During this period, 646,881 patients visited the ED. We use two daily-level data sets from Southlake RHC: Daily Access Reporting Tool (DART) and staff schedules. The DART data set includes two types of variables: service performance and patient volume. For service performance, DART contains 90*^th^* percentiles of TPIA, LOS*^Act NAd^*, LOS*^NAct NAd^*, LOS*^Ad^* and the triage-to-admit duration (T2A). The patient volume measures include (1) total ED visits by CTAS score, (2) total number of admitted patients (ADM) by CTAS, (3) number of patients classified as “alternative level of care” (ALC), and (4) LWBS. We aggregate the ED visits into two new variables: *Acute*, defined as the sum of all CTAS 1, 2 or 3 patients, and *NonAcute* –the sum of CTAS 4 or 5 patients.

While the DART data set includes observations from June 1*^st^*, 2009 to June 2*^nd^*, 2016, the observations for LOS*^Act NAd^*, LOS*^NAct NAd^*, and LOS*^Ad^* only started on March 25*^th^*, 2011, resulting in fewer observations (only 73 prior to the project start) for these variables, which affected some of the analytical choices, as described below.

The staff schedule data contains shift-level physician schedules from January 1*^st^*, 2009 to February 28*^th^*, 2016; from February 27*^th^*, 2013 it also includes navigator schedules. Since our analysis is performed at the daily level, we aggregate staffing information, creating several variables: (1) *DayPhysician*, defined as the total physician-hours during daytime (6 AM - 9 PM), and (2) *NightPhysician*, total physician-hours during night time (9 PM - 6 AM).We use *Staffing Controls* to represent the set of physician-related variables described above.

We make two notes with respect to the staffing data. First, our data set did not include nurse staffing schedules. However, in repeated conversations with SouthLake personnel we were assured that total nurse staffing is (a) constant on each day, and (b) did not change at all during the timeline of the study. Second, the variables defined above for physicians could also be computed for navigators. However, after the introduction of navigators on February 27*^th^*, 2013 it became normal for each physician to be accompanied by a navigator, thus navigator staffing is nearly perfectly correlated with the physician staffing variables described above after that date. Thus, we capture the possible effect of the introduction of navigators with *Nav Use* and *Aft Nav* variables described below.

The next, and, arguably most important, set of variables we create, are variables representing the re-design project, which we call *Project Controls*. These include: (1) a binary variable *Project* with the value of 0 prior to June 6*^th^*, 2011 (the starting day of the project) and value of 1 thereafter, (2) *Aft Project*, which counts the number of elapsed days the project was initiated and takes the value of 0 prior to June 6*^th^*, 2011, (3) a binary variable *Nav Use* with value of 0 prior to February 27*^th^*, 2013 (the starting day of the use of Navigators by SouthLake) and value of 1 thereafter, and (4) *Aft Nav* - the counter of elapsed days from the start of Navigator use, with the value of 0 prior to that date. In addition, we use logarithmic transformations of *Aft Project* and *Aft Nav* to capture possible logarithmic trends (the log-transformed variables are set to the value of 0.0001 when the underlying variable equals to 0).

We also used the date identifier to generate day-of-week and month indicators, as well as the elapsed days counter (i.e., the number of days past since the first observation in the data set - this is to capture general trends). We refer to this group of date-related variables as *Time Controls*. Table 2 provides summary statistics for all the key variables; for each variable the corresponding statistic is computed *before* the project start, and *after* the project start; the overall value in the dataset is also provided.

### Estimation Procedure

Our primary interest is to study the project impacts on ED service performance and patient volume. As noted earlier, we use simultaneous equations (SEMs) as our primary modeling technique. The reasons for using this technique are two-fold. First, key variables in the data such as service performance and patient volume, are likely interacting with each other over time. For example, patient volume and waiting times KPIs are presumably negatively correlated, as higher patient volume induces longer waits, while longer waits in turn attract less patients. SEM is an appropriate tool to capture such interdependent relations among variables. Second, since the dependent variables of any two equations in SEM may be correlated, we expect the error terms of respective equations to be correlated as well. Therefore, we use simultaneous estimation to increase statistical efficiency (Greene 2007).

We develop three sets of equations, each focused on the key sub-task of the estimation procedure. The first SEM is focused on estimating the interrelationships between the TPIA and the patient volume inflow indicators: Acute, Non-Acute, and ADM (Admitted).

We assume that TPIA on day *t* (i.e., *TPIA_t_*) may be affected by the inflows *Acute_t_, NonAcute_t_*. Moreover, since the handling of each patient type changed significantly after the project started, we add the interaction terms *Project_t_* ×*Acute_t_* and *Project_t_* ×*NonAcute_t_*, where *Project_t_* is the binary variable described earlier indicating whether day *t* is before or after the project start. Note that the patient inflow effects are contemporaneous. We also include the number of admissions from the previous day (*ADM_t_*−_1_), since admitted patients frequently spend more than 24 hours in the ED (see Table 2) and thus the previous day’s admission would still impact the processing of new patients on the following day. Finally, we include all Project, Staffing and Time Control variables described earlier. This results in equation (1).

To model the direct impact of reductions in TPIA and the project variables on patient inflows, we assume that the incoming patients (or ambulance dispatchers) can track the TPIA trends at Southlake, however, since the waiting times are not posted, these effects may involve some delays. We therefore form Moving Averages of TPIA values over the previous *t* = 7, 30, and 100 days (not including the current day) and use the resulting variables *TPIA_t-7_^MA^, TPIA_t-30_^MA^* and *TPIA_t-100_^MA^* as predictors in equations (2, 3) for acute and non-acute patient inflows, respectively. Other terms in these equations include Project, Staffing, and Time Controls. Note that the latter allows the model to represent any long-term trends in acute and non-acute patient volumes (e.g., due to population changes in Southlake catchment area).

The last equation in the first SEM is for Admitted patients. The predictors are the number of Acute patients on that day (since admitted patients nearly always arrive with triage scores in the acute category), as well as all Project, Staffing and Time Controls; see equation (4).

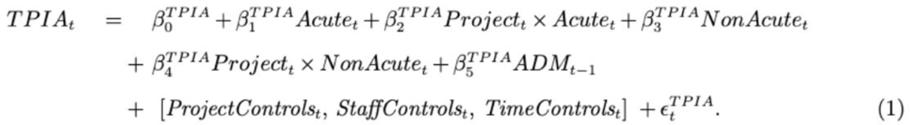

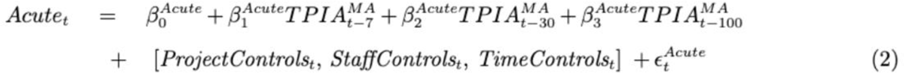

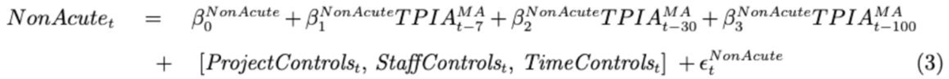

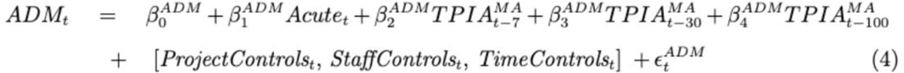

Observe that equations (1-4) allow us to model feedback loops between TPIA and patient inflows: the former is affected by the contemporaneous inflows of Acute and Non-Acute patients, while the equations for the latter are influenced by TPIA through the moving average terms. Whether the feedback is positive or negative is determined by the relative sizes of the corresponding coefficients.

The second SEM is focused on estimating the LOS components, as well as the T2A: 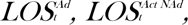 *LOS^NAct Nad^,T*2*A*. Note that these measures are all downstream of TPIA (see Figure 1) and thus are likely to be affected by the TPIA, rather than affect it. The equations have the same structure: the predictors include *TPIA_t_* and its logarithm, the patient inflows *Acute_t_* and *NonAcute_t_*, as well as their interactions with *Project_t_* variable, the number of admitted patients from the previous day *ADM_t_*−_1_, as well as the Project, Staffing and Time Controls. The included predictors can be seen in Table 4.

These equations allow us to capture both linear and non-linear temporal patterns through the inclusion of linear, logarithmic and interaction terms. They also allow us to capture the intercorrelations between the error terms (e.g., larger LOS for one patient group may be correlated with the contemporaneous increases for the other groups). On the other hand, the feedback loops are limited to the first SEM.

It may reasonably be asked why the two separate SEMs are required, i.e., why could all equations not be included in one joint SEM? The main reason is that, as described earlier, our dataset for LOS and T2A measures is substantially shorter than for measures in the first SEM. Moreover, this shorter data starts only 73 days prior to the project start, which may not allow the model to adequately compare the “before” and “after” period. Thus, we rely on the first SEM - which has larger data support (nearly two years prior to the project start and 5 years after) - to model the project impacts on the TPIA and patient inflows (Hypotheses 1 and 2), while using the second SEM to model the impacts on LOS and T2A (Hypothesis 3).

In addition to the two SEMs above, we also construct a separate equation for the LWBS measure. Since this measure represents patient counts with typically low values (as per Table 2, the median daily values are 6 before the project and 2 after), we model it using Poisson regression. This model allows us to analyze Hypothesis 4. In terms of the included predictors, this equation is similar to the other equations in the second SEM, as can be seen from the coefficients in Table 4.

We note some methodological issues related to the estimation of the SEMs. First, the error terms in different equations are correlated. Second and particularly, by analyzing each equation separately, we find that the error term in any equation could either be auto-correlated, or heteroscedastic, or both. This significantly complicates the estimation of the error covariance matrix of two SEMs and prohibits us from using stylized methods such as 3SLS (Greene 2007). To tackle the issue, we adopt the Multi-Stage Least Square (MSLS) method pioneered by LeMay (1990).

Finally, we note that while all the wait time measures in our data (i.e., TPIA, LOS, T2A) were at the daily 90*^th^* percentile level, we are NOT using 90*^th^* quantile regression (since we do not have estimates at other quantiles), but rather the usual mean-level regressions (i.e., we treat the 90*^th^* percentile measurement as simply the measurement protocol of our response variables).

### Iterative Counterfactual Analysis and Bootstrap Error Bounds

Our primary goal is to evaluate the impact of the redesign project on various KPIs and patient inflow measures described above. The question we ask is a *counterfactual*: what would the values of the measures be at each post-project time period had the project never taken place?

In classical regression modeling this question could perhaps be answered by looking at the sign and the significance of the *Project* variable in our model - e.g., a negative and statistically significant coefficient of this variable in the *TPIA_t_* equation would indicate that the project did reduce TPIA.

However, our modeling system described in the previous section is quite complex and is designed to capture both immediate changes in a particular variable (“direct” effects) resulting from the project, as well as “indirect” effects (due to feedback loops). Moreover, our Project Controls is a set of variables designed to represent a pattern of linear or non-linear effects that may evolve over time, and that may be reflected in various projected-related, navigator-related, and interaction terms. As discussed in the following section, this results in a complex pattern of positive and negative coefficients, some of them acting on a linear scale, while others are on the logarithmic scale. Thus, a simple examination of signs and significance of particular terms is not fruitful.

Instead, we introduce a more systematic approach for estimating the trajectory of the system if the project had not taken place: an *iterative counterfactual analysis*. First, we set all project-related variables in each equation to zero. Then, starting with the project start day (i.e., June 6*^th^*, 2011), we compute the estimated values of all endogenous variables in the temporal order of their interaction, i.e., {*Acute_t_* and *NonAcute_t_*} → *ADM_t_* → *TPIA_t_* →{*LOS_t_* measures, *T*2*A_t_, LWBS_t_*}. We then feed these updated values into equations for the next day *t*+1, and so on, until the last day in the dataset. See Figure 2 for the illustration.

**Figure 2.**
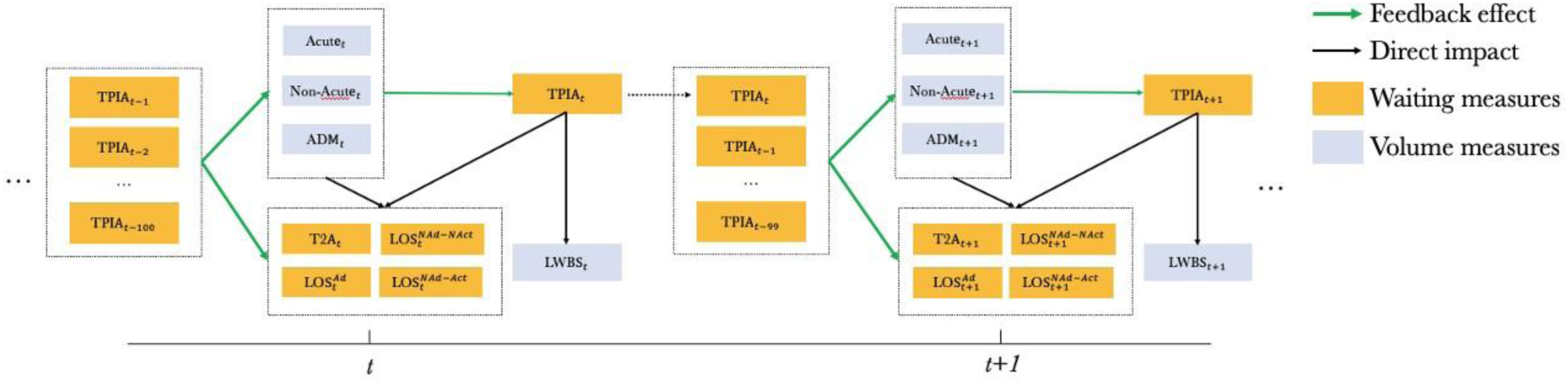
Illustration of Iterative Counterfactual Analysis.

The above procedure yields the *estimated system trajectory under the “no project” condition*, i.e., the *counterfactual*. We then re-apply the same procedure, this time keeping all project controls at their actual values. This yields the *estimated factual trajectory*. Subtracting the latter from the former we obtain the trajectory of *estimated impacts* of the project. These trajectories for each variable are plotted in Figures 3, 4 below. Note that a positive value for a wait-time KPI (i.e., TPIA, LOS, T2A) indicates that the corresponding KPI would have been *higher* under the “no project” condition, i.e., that the KPI improved as a result of the project. For KPIs representing patient counts (Acute, Non-Acute, ADM) the meaning is the opposite: a negative value indicates that the corresponding patient inflow would have been *lower* under the “no project” condition, indicating increase in the corresponding inflow under the project.

**Figure 3.**
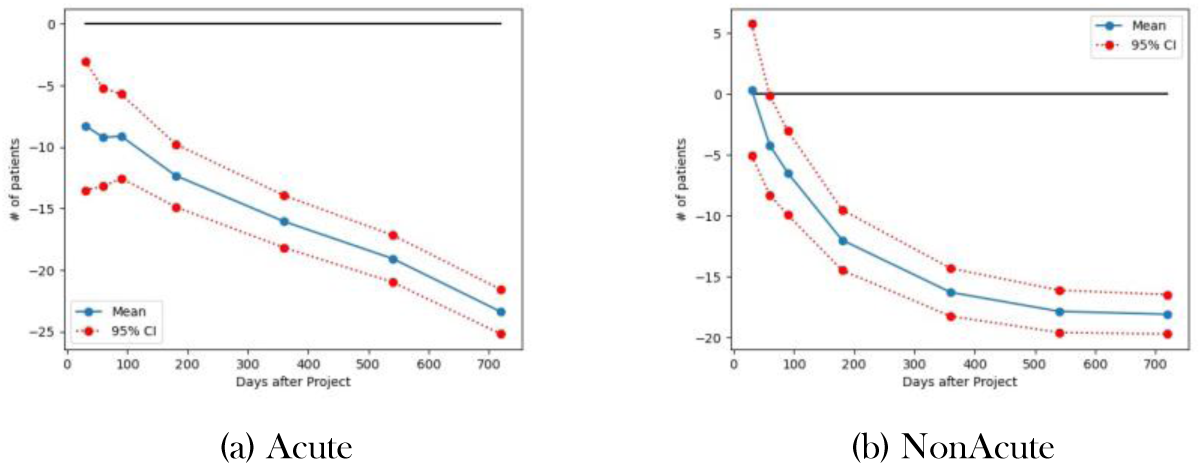
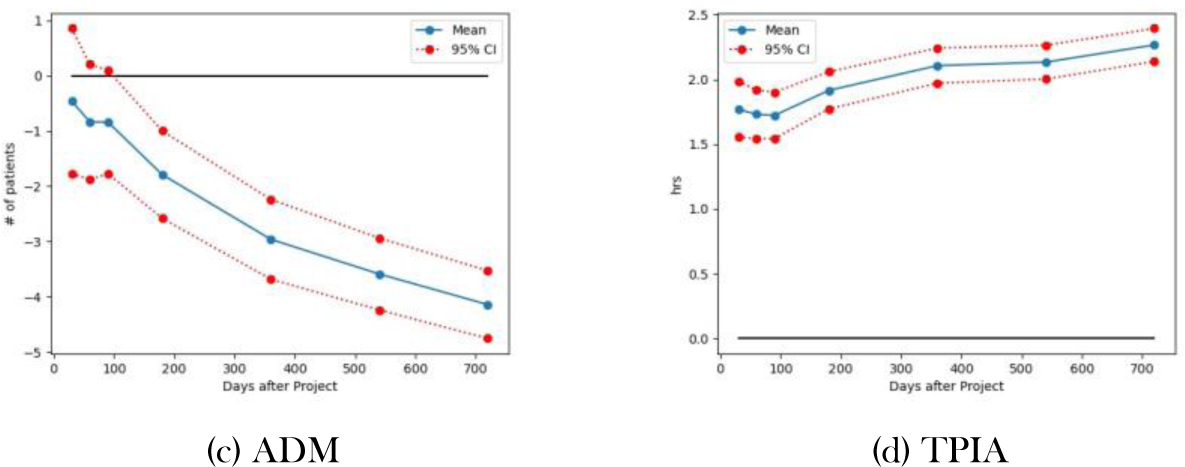
Counterfactual Analysis of SEM 1 (Acute, NonAcute, ADM, TPIA). Each plot represents the estimated difference between the “no project” condition (the counterfactual) and the “with project” condition (the factual).

**Figure 4.**
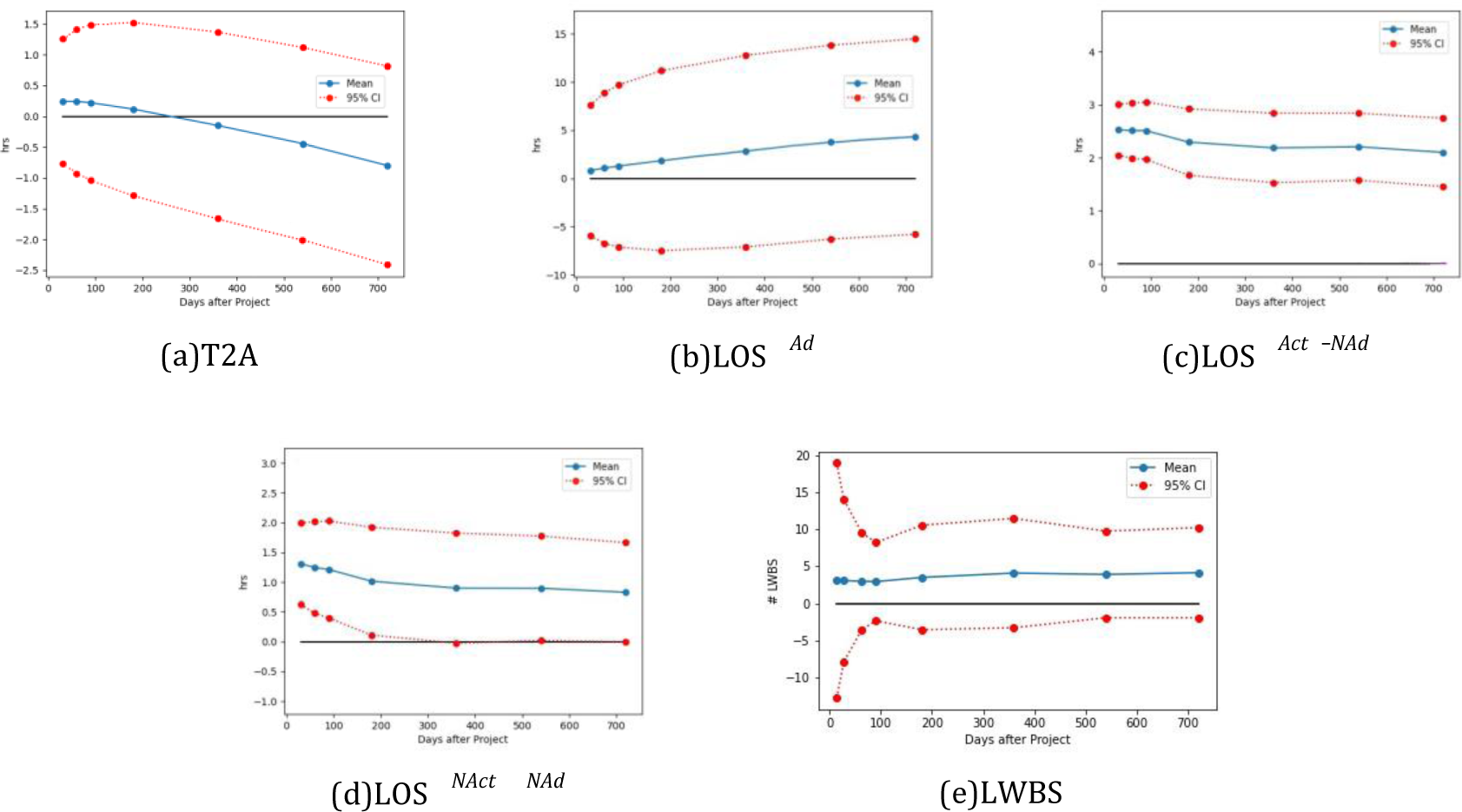
Counterfactual Analysis of Other Service Performance Measures & LWBS. Each plot represents the estimated difference between the “no project” condition (the counterfactual) and the “with project” condition (the factual).

The above procedure focuses on estimating the mean project impacts. To see whether such impacts are statistically significant, we further develop a Bootstrap-based method to compute the standard errors and construct 95% confidence intervals for each estimated mean level impact.

## Results

### Simultaneous Estimation

The estimation results for the first SEM are shown in table 3. Table 4 contains results for the second SEM and LWBS.

**Table 3.**
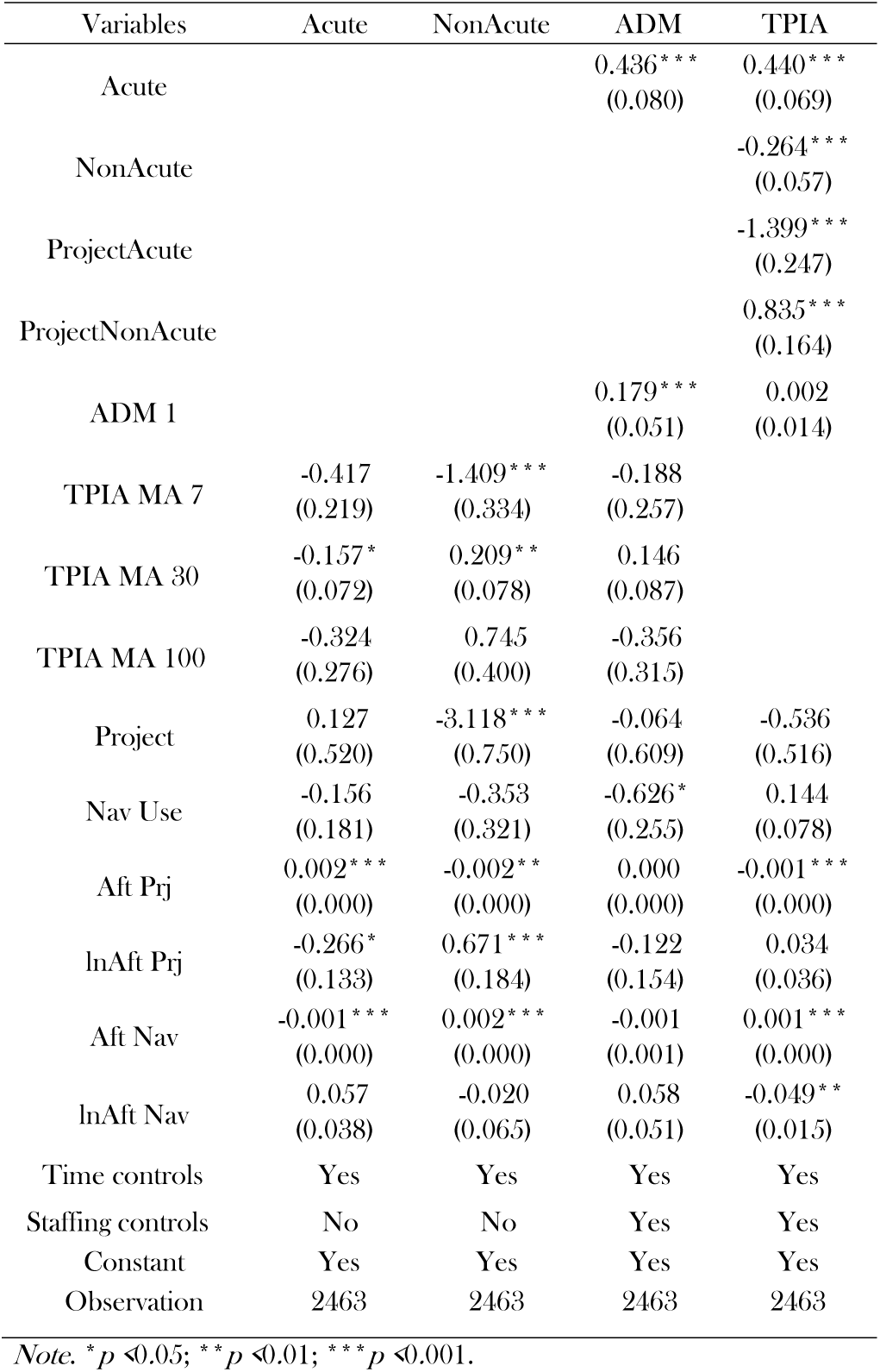
MSLS estimation results for the first SEM (equations 1-4)

**Table 4.**
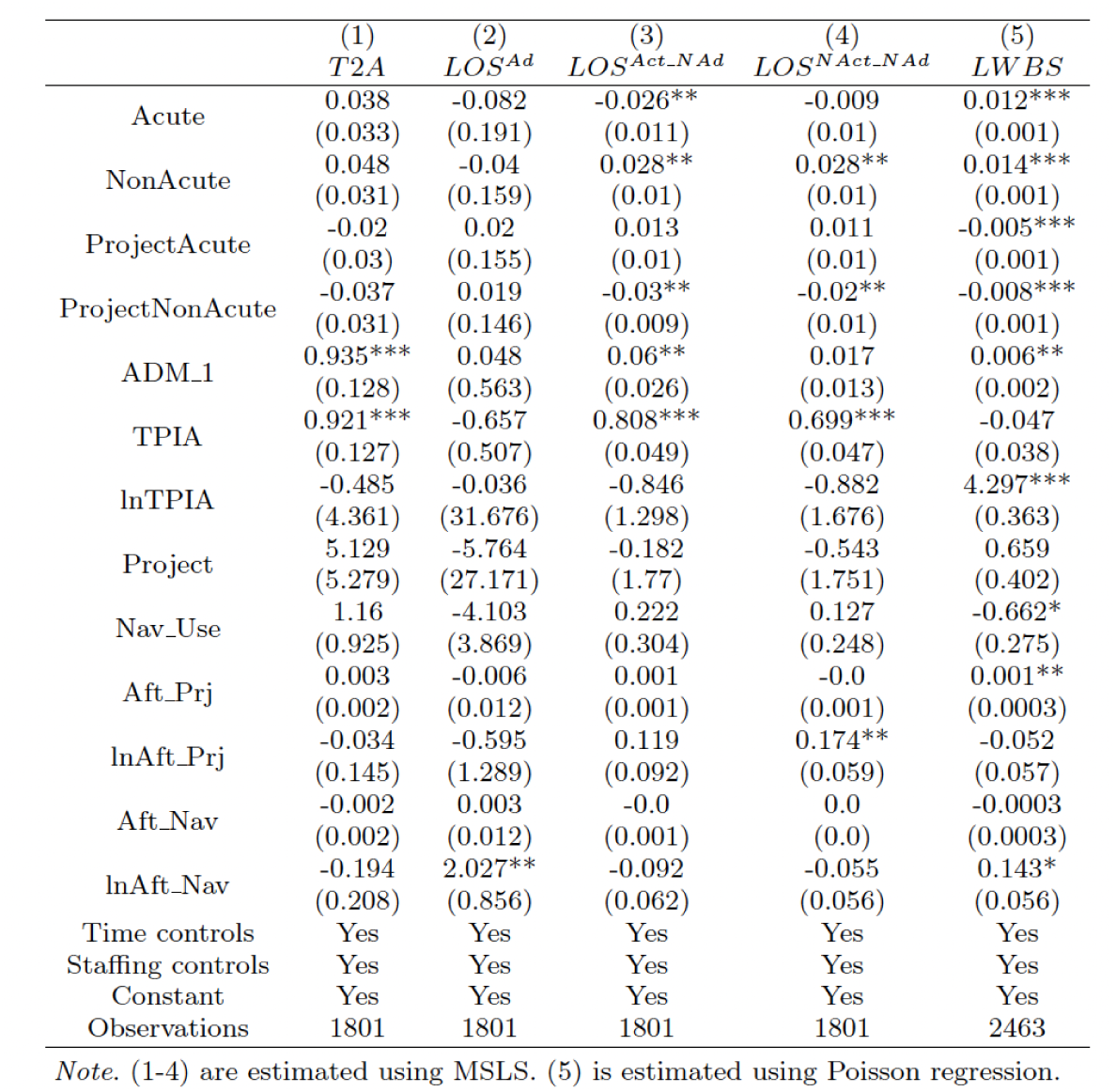
Estimation results for the Second SEM and LWBS.

As noted earlier, these results are difficult to interpret directly since the coefficients of various project controls and endogenous covariates are pointing into conflicting directions. For example, consider the TPIA equation in Table 3. Looking only at significant coefficients (designated by “*”), we see a negative coefficient in front of *Aft Prj*, which represents the number of days since the start of the project, and indicates a linear decrease in TPIA. On the other hand, we see a positive term in front of *Aft Nav* (number of days since the first use of navigators) is positive, indicating a counter-intuitive increase in TPIA, though since this coefficient is applied to a smaller value (since the use of navigators started some time after the project starts), it may just indicate a decrease in the rate of TPIA improvement in more recent periods. A significant negative coefficient in front of *lnAftNav* indicates a non-linear effect. The project also enters into the interaction terms for Acute and Non-Acute patient counts, with different coefficients. In short, the direction and the size of the overall project impact are nearly impossible to determine without the counterfactual analysis that is discussed below. The same consideration applies to other equations in Tables 3 and 4.

### Counterfactual analysis

Figures 3 and 4 present the estimated project impacts with 95% confidence intervals (CI) for every key variable over 720 days post project start. As noted earlier the impact at a given number of days after the project starts is the difference between the counterfactual estimate and the factual (“with project”) estimate for each variable.

Figure 3(d) represents the results for TPIA and *confirms our hypothesis* **H1**: over time, the redesign project has significantly improved the 90*^th^* percentile of TPIA, which was the primary goal of the project. The plot shows both an immediate reduction of about 1.75 hours and the long-run continuous improvement, leading to the ultimate reduction of over 2 hours at 720 days after the project. Relative to the pre-project level of 4.1 hours, this represents an over 50% improvement (after accounting for the impact of all other variables in the model). It is interesting to note that the improvement rate does vary over time: after the immediate initial reduction, there appears to have been a small dip in performance, which then recovered about 100 days after the project started. The differences in improvement rates likely correspond to various continuous improvement initiatives undertaken by Southlake after the start of the project.

With respect to our hypothesis **H2** (that the redesign project led to increased ED patient inflows), from figures 3(a) and (b), we see that the curves for both Acute and Non-Acute arrivals are in the negative region, with the 95% confidence intervals also below zero. This indicates that without the project, both types of patient inflows would have been significantly lower, indicating that the project led to the increase in patient inflows, *thus confirming hypothesis H2*.

For Acute patients, this increase (accounting for all other factors in the model) started at around 10 patients per day 100 days after the project started and continued to grow at a nearly linear rate to the value of around 22 additional patients per day at the 720-day mark. This represents a 23% increase compared to the pre-project level of 137 Acute arrivals per day.

For Non-Acute patients we also observe an increase in the number of visits post-project (after accounting for all other factors), however the pattern is quite different: after an initial steep rate of increase, the growth in Non-Acute arrivals has flattened out at around 540 days post-project, settling at around 18 additional Non-Acute arrivals per day (about 25% increase over the pre-project average of 70).

Not surprisingly, the increase in patient arrivals (particularly Acute arrivals) led to a significant increase in the number of Admissions, as evidenced by Figure 3(c). It is interesting that this curve appears to be flattening out over time, while the rate of growth in Acute arrivals on panel (a) is increasing. There may be several reasons for this effect, ranging from a large part of new Acute arrivals being less urgent patients not requiring a hospital admission, to more efficient treatment procedures (e.g., the introduction of navigators) providing speedier treatment and diverting some potential admissions. Further examination of this interesting effect is needed.

Our hypothesis **H3**, that the redesign project did not lead to deteriorating performance in any of the LOS measures, *is confirmed* by Figure 4, panels (b),(c) and (d). We observe that the blue curve is positive and fairly stable over time, indicating that the corresponding measures improved after the project (i.e., they would have been higher under the counterfactual condition). For admitted patients (panel b) this improvement is not statistically significant, as the confidence intervals span 0. For Acute Non-Admitted patients, the improvement is statistically significant and represents a decrease of about 2.5 hours in LOS, once other factors (including increased patient inflows) are accounted for. For the Non-Acute Non-Admitted group, the improvement is about 1 hr of LOS and is borderline-significant. Finally, we note that the Time to Admission Decision (T2A) measure does not appear to have been significantly affected by the project - see panel (a) - though the downward trend of the mean-level difference curve may represent a trend towards longer decision times.

Our final hypothesis **H4**, that the project led to a decrease in LWBS, *is not supported by the data*. This is evident from panel (e) of Figure 4. While the blue line is positive and steady at between 3 and 4 (indicating an average decrease in LWBS by 3 to 4 patients per day after the project was implemented), the confidence interval lines span 0. Thus, while the decrease in LWBS is observed, it is not statistically significant. This is likely due to the low LWBS counts in the data, requiring a longer sample to establish statistical significance.

## Discussion

Our analysis and results are of both methodological and clinical significance. With respect to the methodology, we demonstrate the importance of taking into account the variety of impacts a large re-design project can have, both cross-sectionally and longitudinally. A simplistic analysis, e.g., *t*−test comparing various before and after measures in Table 2, is not sufficient to account for the temporal evolution of the system or possible feedback. This is especially true in case of large system-wide projects that include multiple stages and use continuous improvement techniques over time. Our methodology, based on SEMs with time-series components supplemented with the iterative counter-factual analysis and bootstrap-based evaluation of statistical significance, is capable of isolating the effects of system re-design on various KPIs and system components. The counterfactual analysis in Figures 3 and 4 shows that the project impacts do evolve over time in a variety of complex patterns.

Arguably, the clinical impacts of our analysis are even more significant. The re-design project achieved a seemingly impossible task: a very large (over 50%) improvement in time to initial assessment (TPIA), without any negative impacts on LOS (in fact, improving LOS for non-admitted Acute and Non-Acute patients), while attracting significantly larger inflows of both acute and nonacute patients. All of this was accomplished without any increase in physician hours (see Table 2).

Note that, in principle, achieving an improvement in TPIA by having the physicians prioritize the initial assessment of the new patients over the reassessments and treatment of the patients already in the ED. However, such a strategy would result in trading off improvement in TPIA vs at least some of the LOS measures. Southlake was clearly aware of this risk and took steps to prevent such trade-offs from happening (see, in particular, the use of “Navigator Routing” on 1). Our finding suggests that these steps were quite successful.

Southlake saw large increases in both acute and non-acute patient inflows after the start of the project. Our findings confirm that these increases can be directly attributed to the TPIA improvements resulting from the project. While we do not directly control for demographic patterns (e.g., we do not have population counts in the model), we control for these confounding effects indirectly by including *time controls* into patient volume equations, which do allow for trends in patient inflows due to exogenous factors. Instead, we find that the driver behind these inflows is the ED redesign project.

This finding supports the anecdotal evidence that the inflows were due to the word-of-mouth (WOM) effects (Martin 2016). Indeed, Tu and Lauer (2008) find from a national study in the U.S. that half of the consumers “relied on word-of-mouth recommendations from friends and relatives doctor recommendations and health plan information” when looking for primary care physicians. Pavlin et al. (2020) fit a discrete choice model using cataract surgical procedure data from Ontario, Canada and conclude that, the wait for a surgical consultation will significantly lower the probability of physician referrals. Anecdotally, there were reports of ambulances being directed to the Southlake hospital, as it provided quicker patient offload, and also of family physicians directing their non-urgent patients to Southlake. Our result provides further evidence of WOM in the ED setting, while also showing the different patterns of effects for Acute and Non-Acute patients.

The fact that despite the significant increase in patient volumes, Southlake was able to maintain improvements in TPIA without compromising LOS measures or increasing the number of physician hours is a testament to the effectiveness of various initiatives undertaken under the redesign project. Indeed, one would expect a deterioration in performance as patient flows increase. Such effects are well-documented in the literature. Indeed, even an increase in just the Non-Acute inflows has been shown to affect waiting times for both Acute and Non-Acute patients - see Bayati et al. (2017).

The project components that likely effectively counteracted these effects include (1) eliminating pre-ED waiting area, allowing physicians to directly observe the waiting patients, which has been documented to service rate and patient flows in ED operations (see Pennathur et al. 2011 and reference therein), (2) not assigning ambulatory patients to a bed, which prevents ED beds from becoming a bottleneck, (3) Navigator preparation work, which increases physician efficiency, (4) scheduling flexibility, which allows the system to automatically adjust capacity to unexpected ebbs and flows of arrivals, and (5) physician capacity leveling, that ensures that the available physician processing capacity matched the expected arrival rate in each shift. We refer to Table 1 and to Whatley (2016) for further description of each initiative.

Overall, the ED staff, aided by the initiatives listed above, managed to greatly increase their efficiency to handle greater patient flows while improving nearly all KPIs. This, of course, required the full buy-in of nursing and physician staff into the redesign project.

We close by noting that this study, like any other, has a number of limitations. First, we analyzed the data at an aggregate (daily) level, which prevents us from studying patient-specific effects requiring patient-level data. Second, the available capacity data (doctor and navigator schedules) reflected the assigned shifts, rather than the actual worked shifts. In particular, we did not have actual arrival/ departure times of doctors of navigators, nor did we have data for on-call doctors who were called in. We had no data for nurse schedules or actual work - we had to rely on assurances that neither the staffing levels nor schedules changed during the project. The fact that our waiting time indicators (TPIA and LOS) were measured at the 90*^th^* percentiles likely made the underlying distributions more skewed, which may reduce the efficiency of the MSLS estimators (though they remain consistent); our bootstrap-based significance estimates should be robust to these effects. Since our analysis seeks to establish a pattern of causal links, an omitted variable bias is always a concern. In particular, the ED is part of a larger system - the Southlake RHC, and is, no doubt, affected by many processes outside of ED. Thus, bringing other system components into our models may have improved accuracy and uncovered some interesting interrelationships, albeit at the cost of further complications to an already fairly complex modeling approach.

## Conclusions

In this paper we study a wide range of longitudinal and cross-sectional impacts of a major and long-term ED re-design project at Southlake RHC. By applying a novel methodology based on simultaneous equation modeling coupled with interactive counterfactual analysis, we are able to identify the effects of the re-design project while accounting for a variety of internal and external factors. We confirm that the system re-design project was indeed able to significantly reduce TPIA over time, without compromising other service performance measures such as T2A or LOS. We also find evidence of the “word-of-mouth” effect, with more patients choosing to attend the hospital as a result of improvements in waiting time KPIs. Our results strongly suggest that other EDs should strongly consider undertaking similar system redesign projects.

Our work can be extended in a number of ways. First, the SEM approach we used is a generalization of regression-based models. Modern machine-learning approaches typically achieve higher accuracy but at the cost of less transparency. The advantage of regression-based approaches is supposed to be the visibility and interpretability of the coefficients. However, as we have seen, the coefficients are, in fact, not very interpretable in a complex setting with multiple equations and time-series terms - we had to recover interpretability through the iterative counter-factual approach. This could be coupled with a machine learning model (such as a neural network or random forest), which may lead to increased accuracy.

On the clinical side, it would be very useful to estimate some of the individual effects of various initiatives in Table 1. This, of course, would require more granular data, both at the patient and physician scheduling level.

## Data Availability

All data produced in the present study are available upon reasonable request to the authors.

